# Comparing between survived and deceased patients with Diabetes Mellitus and COVID-19 in Bangladesh: A cross sectional study from a COVID-19 dedicated hospital

**DOI:** 10.1101/2021.04.04.21254884

**Authors:** Md. Shahed Morshed, Abdullah Al Mosabbir, Mohammad Sorowar Hossain

## Abstract

The current coronavirus disease 2019 (COVID-19) outbreak was reported to cause significantly higher mortality and morbidity among patients with diabetes mellitus (DM). Although Bangladesh is amongst the top 10 countries with diabetic people, data on these patients with COVID-19 is scarce from this region. This study aimed to illustrate the clinical features and outcomes of hospitalized patients with COVID-19 and DM in Bangladesh while comparing survivors and deceased.

This retrospective cross-sectional study was conducted among RT-PCR confirmed COVID-19 patients with pre-existing Diabetes Mellitus in a specialized COVID-19 hospital in Bangladesh. Data from hospital records were analyzed.

Among 921 RT-PCR confirmed COVID-19 admitted during the study period, 231 (∼25%) patients with pre-existing DM (median age 60 years) were included in the analysis. The death rate among all hospitalized patients (with and without DM) was 2.8% compared to 11.3% among diabetic patients. The median hospital stay was 13 days (IQR 10.5, 17.0) for survivors and five days (IQR 2.0-8.3) for the deceased. The clinical features were not significantly different between survivors and the deceased. However, deceased patients had significantly lower blood oxygen level (85% vs 93%, p <0.001), and higher neutrophil-lymphocyte ratio (7.9 vs 4.5, p 0.003) and serum ferritin (946.0 vs 425.0 ng/ml, p 0.03). Glycemic status was poor in both groups.

This study would help identify a subgroup of diabetic patients with COVID-19 who are at higher risk of in-hospital death and improve clinical decision making.

## Introduction

The ongoing pandemic of coronavirus disease 2019 (COVID-19) is one of the greatest disasters that the world has ever witnessed. While the clinical spectrum of COVID-19 is highly variable, the disease severity and mortality were reported to be significantly higher among patients with comorbidities, particularly diabetes mellitus (DM) [1,2]. India, Pakistan, and Bangladesh are among the top 10 countries globally by the number of affected people and undiagnosed cases [3]. Despite diabetes being a major public health concern, there are limited studies on diabetic patients with COVID-19 from these regions, especially Bangladesh. Moreover, studies comparing survived and diseased patients with DM and COVID-19 are rare in literature, which we believe is vital to identify cases requiring urgent clinical attention.This study aimed to address the clinical epidemiology and outcome of COVID-19 patients with DM in Bangladesh, along with a detailed comparison between survived and deceased.

## Methods

This retrospective cross-sectional observational study was conducted among RT-PCR confirmed COVID-19 patients with pre-existing DM admitted from 1 to 30 June 2020 in a specialized COVID-19 hospital (Kurmitola General Hospital) located in Dhaka, the epicenter of the COVID-19 pandemic in Bangladesh. Patients with known DM and taking antidiabetic agents were considered to have pre-existing DM. Patients having glycated hemoglobin (HbA1c) _≥_6.5% done within three months of admission were also included as having pre-existing DM. The severity of COVID-19 was described by WHO interim guidance [4]. Data were extracted from hospital records using a relevant questionnaire. All information was double-checked before analysis to ensure quality. The institutional review board of the Biomedical Research Foundation, Bangladesh, approved the study protocol (Ref. no: BRF/ERB/2020/003). Data were analyzed by SPSS 22.0 software.

## Results

A total of 921 RT-PCR confirmed COVID-19 patients were admitted to the hospital during the study period. Of them, 231 (25.0%) patients with pre-existing DM were included in this study. While the death rate among all hospitalized patients (with and without DM) was only 2.8% (58/921), it was four times higher (11.3%, 26/231) among patients with DM (Table 1). The median age of the deceased patients with DM was slightly higher compared to survived (63.5 vs 59 years, p 0.21). The most common comorbidity in both survived and deceased patients were hypertension (HTN) (59.5% and 73.1%, respectively).The clinical presentations were not significantly different between survived and deceased except for significantly lower oxygen saturation among deceased (85% vs 93%, p <0.001). The absolute neutrophil count and neutrophil-lymphocyte ratio (NLR) of deceased patients with DM were significantly higher than survived. Besides, serum ferritin was more than two times higher among the deceased (946.0 vs 425.0 ng/ml, p 0.03). The glycemic status was poor in both survived and deceased patients; median HbA1c was 8.3% and 8.6%, respectively. In addition, random blood glucose (RBG) at admission was significantly higher among deceased (16.9 vs 14.0 mmol/L, p 0.002). Overall, the median hospital stay for patients with DM was 12 days (IQR 10.0-16.0). Interestingly, hospital days were significantly higher among survivors (13 vs 5 days, p <0.001). While intensive care unit (ICU) support was indicated for 31.2% (72/231) patients with DM, only 12.5% (29/231) of them got ICU admission (Fig 1).The mortality rate among ICU patients with DM was 46% (12/29). Among antidiabetic agents used, a significantly higher proportion of deceased patients required insulin than survived (Table S1).

**Table 1:** Baseline characteristics, clinical features and laboratory findings of patients with COVID-19 and DM^1, 2^

| **Variables** | **Total** | **Survived** | **Deceased** | **P value** |
| --- | --- | --- | --- | --- |
| ***Baseline Characteristics*** | 231 (100) | 205 (88.7) | 26 (11.3) |  |
| Age, years | 60.0 (51.0-65.0) | 59.0 (51.0-65.0) | 63.5 (50.0-68.3) | 0.21 |
| Age groups |  |  |  |  |
| <40 | 17 (7.4) | 14 (6.8) | 3 (11.5) | 0.05 |
| 40 – 59 | 96 (41.6) | 91 (44.4) | 5 (19.2) |  |
| ≥60 | 118 (51.1) | 100 (48.8) | 18 (69.2) |  |
| Sex |  |  |  |  |
| Male | 151 (65.4) | 132 (64.4) | 19 (73.1) | 0.40 |
| Female | 80 (34.6) | 73 (35.6) | 7 (26.9) |  |
| Comorbidities<sup>3</sup> |  |  |  |  |
| Any | 174 (75.3) | 150 (73.2) | 24 (92.3) | **0.05** |
| Hypertension | 141 (61.0) | 122 (59.5) | 19 (73.1) | 0.21 |
| Chronic Kidney Disease | 41 (17.7) | 34 (16.6) | 7 (26.9) | 0.27 |
| Ischemic Heart Disease | 36 (15.6) | 28 (13.7) | 8 (30.8) | **0.04** |
| Obstructive lung disease | 22 (9.5) | 18 (8.8) | 4 (15.4) | 0.29 |
| Cerebrovascular disease | 11 (4.8) | 9 (4.4) | 2 (7.7) | 0.36 |
| ***Clinical features***<sup>4</sup> |  |  |  |  |
| Predominant symptoms |  |  |  |  |
| Fever | 164 (71.0) | 146 (71.2) | 18 (69.2) | 1.00 |
| Breathlessness | 149 (64.5) | 129 (62.9) | 20 (76.9) | 0.20 |
| Cough | 132 (57.1) | 122 (59.5) | 10 (38.5) | 0.06 |
| Anorexia | 20 (8.7) | 19 (9.3) | 1 (3.8) | 0.71 |
| Sore throat | 15 (6.5) | 15 (7.3) | 0 (0.0) | 0.23 |
| Signs at presentation |  |  |  |  |
| Temperature (°C) | 36.0 (35.6-36.4) | 36.0 (35.6-36.4) | 36.2 (35.8-36.7) | 0.10 |
| Oxygen saturation (%) | 93.0 (86.0-96.0) | 93.0 (88.0-96.0) | 85.0 (79.0-91.0) | **<0.001** |
| Severity of disease at presentation |  |  |  |  |
| Mild | 78 (33.8) | 75 (36.6) | 3 (11.5) | **<0.001** |
| Moderate | 68 (29.4) | 63 (30.7) | 5 (19.2) |  |
| Severe | 83 (35.9) | 67 (32.7) | 16 (61.5) |  |
| Critical | 2 (0.9) | 0 (0.0) | 2 (7.7) |  |
| Hospital days | 12.0 (10.0-16.0) | 13.0 (10.5-17.0) | 5.0 (2.0-8.3) | **<0.001** |
| ***Laboratory Findings***<sup>5</sup> |  |  |  |  |
| Hemoglobin, gm/dL | 11.8 (10.5-13.3)<br>[185] | 11.8 (10.7-13.3)<br>[169] | 10.8 (9.6-13.2)<br>[16] | 0.24 |
| Total leukocytes, ×10<sup>3</sup>/μL | 8.6 (6.8-12.8)<br>[185] | 8.4 (6.6-11.3)<br>[169] | 14.8 (10.9-19.1)<br>[16] | **<0.001** |
| Absolute neutrophils count, ×10<sup>3</sup>/μL | 6.61 (4.54-10.34)<br>[185] | 6.25 (4.49-9.34)<br>[169] | 13.11 (9.31, 15.83) [16] | **<0.001** |
| Absolute lymphocytes count, ×10<sup>3</sup>/μL | 1.55 (1.05-2.16)<br>[185] | 1.55 (1.05-2.15)<br>[169] | 1.55 (1.05-2.24)<br>[16] | 0.969 |
| Neutrophils:Lymphocytes | 4.6 (2.6-7.7)<br>[185] | 4.5 (2.4-7.1) [169] | 7.9 (4.3-10.0) [16] | **0.003** |
| Platelet count, $\times 10^3/\mu\text{L}$ | 261.0 (185.0-354.0) [185] | 256.0 (181.0-350.5) [169] | 289.5 (212.0-364.8) [16] | 0.58 |
| Alanine aminotransferase, U/L | 54.5 (36.5-87.3) [154] | 54.0 (35.0-86.0) [139] | 62.0 (53.0-88.0) [15] | 0.34 |
| Serum creatinine, mg/dL | 1.2 (1.0-1.5) [178] | 1.1 (1.0-1.4) [159] | 1.2 (1.0-1.9) [19] | 0.440 |
| eGFR ml/min/1.73 m<sup>2</sup> | 62.50 (43.25-78.25) [178] | 63.0 (45.0-79.0) [159] | 56.0 (34.0-70.0) [19] | 0.435 |
| Serum Sodium, mmol/L | 139.0 (135.0-142.0) [163] | 139.0 (136.0-142.0) [142] | 137.0 (128.5-141.0) [21] | 0.130 |
| Serum Potassium, mmol/L | 4.4 (4.0-4.8) [163] | 4.4 (4.0-4.9) [142] | 4.2 [3.9-4.8] [21] | 0.40 |
| Serum C-reactive protein, mg/L | 24.0 (6.0-24.0) [156] | 24.0 (6.0-24.0) [140] | 24.0 [7.5-24.0] [21] | 0.40 |
| Serum ferritin. ng/ml | 463.5 (226.0-987.3) [96] | 425.0 (213.0-936.0) [83] | 946.0 (399.5-1690.5) [13] | **0.03** |
| Serum D-dimer, mg/L | 0.4 (0.2-1.1) [97] | 0.4 (0.1-1.1) [88] | 0.6 (0.3-6.4) [9] | 0.13 |
| Serum lactate dehydrogenase, U/L | 507.5 (375.0-683.0) [44] | 480.0 (373.0-640.0) [39] | 750.0 (461.0-958.0) [5] | 0.15 |
| Glycemic profile |  |  |  |  |
| Random blood glucose, mmol/L | 14.9 (9.6-18.2) [231] | 14.0 (9.1-18.2) [205] | 16.9 (15.3-20.0) [26] | **0.002** |
| HbA1c (%) | 8.3 (7.1-9.5) [50] | 8.3 (7.3-9.5) [41] | 8.6 (6.8-10.0) [9] | 0.90 |

<sup>1</sup>Pearson's chi-square/ Fisher's exact test or Mann-Whitney U test was done as appropriate; p-value <0.05 was set as statistically significant
<sup>2</sup>Number (%) or Median (IQR) was used where appropriate; Parentheses: (percentages over column total of each variable); [available number]
<sup>3</sup>Other comorbidities: hypothyroidism (6), chronic liver disease (3), benign enlargement of prostate (2), malignancy (1), rheumatoid arthritis (1), gout (1), etc
<sup>4</sup>Other symptoms: fatigue (14), diarrhea (11), vomiting (10), chest pain/ tightness (7), headache (5), altered taste/ smell (5), dizziness (3), etc
<sup>5</sup>Data were not available for all patients as investigations were done on a need basis

**Figure 1:**
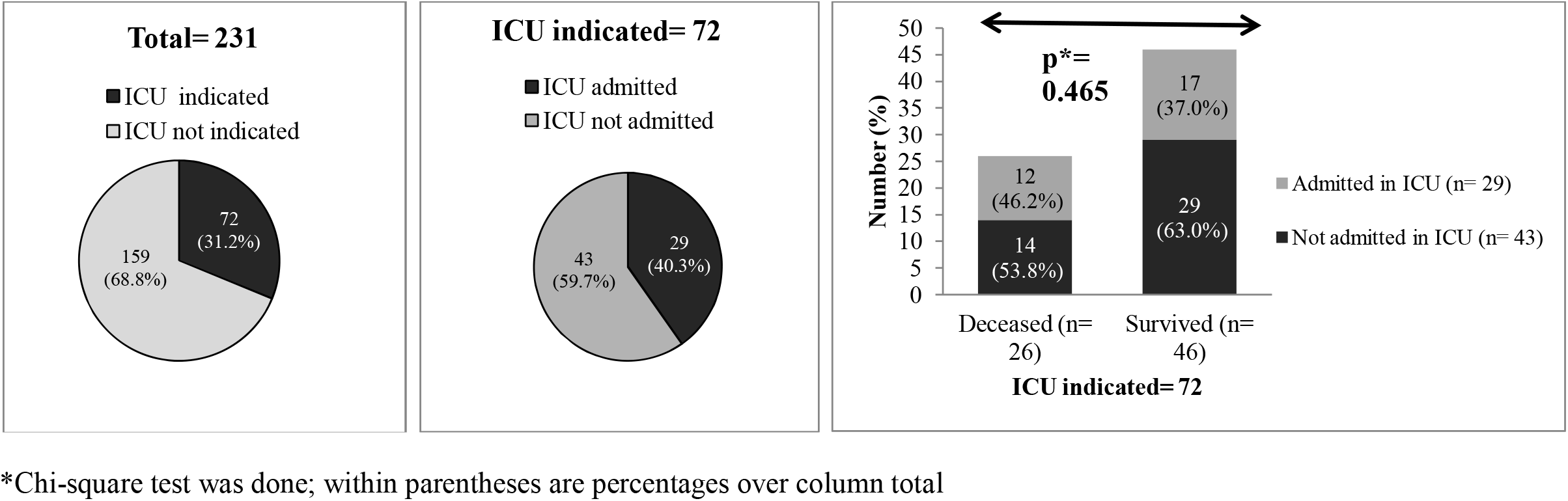
Outcome of patients with DM and COVID-19 who required ICU care. Patients referred to ICU with proper indication were considered as ICU care indicated.

## Discussion

Our study found that one in four hospitalized patients (235/921) with COVID-19 had pre-existing DM. One study from the southern region of Bangladesh reported a slightly lower prevalence (19.8%) of DM among hospitalized patients [5]. The prevalence showed wide variability in other countries ranging from 5.3% in China to 33% in Europe and nearly 50% in India [1,6]. This could be due to regional variation of healthcare policy and perception of diabetic patients towards COVID-19. In this study, the mortality rate among hospitalized patients with DM was more than 11%. However, the overall case fatality rate in Bangladesh is only 1.5% [7]. This trend largely agrees with other studies, althoughthe death rate was comparatively higher in our study [1,6,8]. While we are not certain about the cause of this observation, we found that some parameters were significantly different between two groups. While both deceased and survived patients with DM had poor glycemic control, deceased patients had significantly higher RBG.The association between poorly controlled blood glucose and higher mortality in patients with DM and COVID-19 has been reported in many studies^1,9^. Additionally, median NLR and serum ferritin levels were almost double among deceased compared to survived. Highly elevated NLR and serum ferritin essentially indicate excessive inflammation leading to cytokine storm and multiple organ damage in patients with COVID-19. Significantly higher NLR and serum ferritin levels among non-survivors compared to survivors were reported in already published studies [10,11].

Our study reports another intriguing observation. While ICU support was indicated in over 30% (72/231) of diabetic patients, more than half of them (43/72) could not be admitted into ICU due to the acute bed crisis (only a ten bedded ICU support in the study hospital). Interestingly, the mortality rate was not significantly different between patients admitted into the ICU and those who were not but required ICU care (46.2% vs 53.2%, p 0.465). We could not find any literature to compare our observations as this kind of situation is unique for countries with limited resources like Bangladesh. We believe this is a unique finding for resource-poor settingsand needs further research. Small sample size and precautious refferal to ICU of relatively less severe patients might be two important factors.

Our study has some limitations. The sample size was not large enough to evaluate the predictive performance of different parameters for death in diabetic patients. Besides, we could not collect data on the type of DM or new-onset DM.

## Conclusions

Our study found deceased patients with DM and COVID-19 had significantly lower oxygen saturation and significantly higher NLR, RBG and serum ferritin levels compared to survived. We believe findings from this study will help clinicians to identify a subgroup of diabetic patients with COVID-19 who are at higher risk of in-hospital death; hence, requiring rigorous clinical management.

## Supporting information

Not applicable

## Data Availability

Not applicable. All data are included in the manuscript

## Acknowledgement

We are grateful to Brigadier General Jamil Ahmad, director, Kurmitola General Hospital (KGH) and all nurses of KGH for their generous support in data collection. We humbly acknowledge the contribution of our patients as well.

## Authors’ contributions

**MSM:** Conceptualization, Methodology, Formal analysis, Data Curation, Writing - Review & Editing; **AAM:** Methodology, Writing - Original Draft, Visualization; **MSH:** Writing - Review & Editing, Supervision

