## Supplementary material for "Comparing between survived and deceased patients with Diabetes Mellitus and COVID-19 in Bangladesh: A cross sectional study from a COVID-19 dedicated hospital": Not applicable

**Table S1:** Treatment of COVID-19 patients with DM (n=231)^1^

| Variables | Total | Survived | Deceased | P value |
| --- | --- | --- | --- | --- |
| Antivirals^2^ | 51 (22.1) | 42 (20.5) | 9 (34.6) | 0.13 |
| Oral antibiotics^3^ | 182 (78.8) | 165 (80.5) | 17 (65.4) | 0.12 |
| Intravenous antibiotics | 154 (66.7) | 136 (66.3) | 18 (69.2) | 0.83 |
| Ivermectin | 147 (63.6) | 136 (66.3) | 11 (42.3) | **0.02** |
| Anticoagulants^4^ | 203 (87.9) | 182 (88.8) | 21 (80.8) | 0.33 |
| Steroids | 129 (55.8) | 113 (55.1) | 16 (61.5) | 0.68 |
| Convalescent plasma therapy | 11 (4.8) | 6 (2.9) | 5 (19.2) | **0.004** |
| Tocilizumab | 9 (3.9) | 5 (2.4) | 4 (15.4) | **0.01** |
| ACEI/ ARB | 122 (52.8) | 107 (52.2) | 15 (57.7) | 0.68 |
| Oral antidiabetic agents | 94 (40.7) | 89 (43.4) | 5 (19.2) | **0.02** |
| Insulin | 161 (69.7) | 138 (67.3) | 23 (88.5) | **0.04** |

ACEI, angiotensin converting enzyme inhibitors; ARB, angiotensin-receptor blockers

^1^Within parentheses are percentages over column total of each variable; Number (%) was used

^2^Antivirals: favipiravir-27 (11.7%); remdesivir- 25 (10.8%)

^3^Oral antibiotics: doxicycline- 122 (52.8%); coamoxiclav- 61 (26.4%); azithromycin- 42 (18.2%), etc

^4^Anticoagulant: Low molecular weight heparin, all enoxaparin
